# The Interaction of Child Abuse and rs1360780 of the FKBP5 Gene is Associated with Amygdala Resting-State Functional Connectivity in Young Adults

**DOI:** 10.1101/2020.11.25.20238519

**Authors:** Christiane Wesarg, Ilya M. Veer, Nicole Y. L. Oei, Laura S. Daedelow, Tristram A. Lett, Tobias Banaschewski, Gareth J. Barker, Arun L.W. Bokde, Erin Burke Quinlan, Sylvane Desrivières, Herta Flor, Antoine Grigis, Hugh Garavan, Rüdiger Brühl, Jean-Luc Martinot, Eric Artiges, Frauke Nees, Dimitri Papadopoulos Orfanos, Luise Poustka, Sarah Hohmann, Juliane H. Fröhner, Michael N. Smolka, Robert Whelan, Gunter Schumann, Andreas Heinz, Henrik Walter, IMAGEN Consortium

**Author notes:** Authors contributed equally (Christiane Wesarg, Ilya M. Veer). Corresponding author: Ilya M. Veer, PhD, Charité - Universitätsmedizin Berlin, Klinik für Psychiatrie und Psychotherapie, Campus Mitte, Charitéplatz 1, 10117 Berlin.

## Abstract

Extensive research has demonstrated that rs1360780, a common single nucleotide polymorphism within the FKBP5 gene, interacts with early-life stress in predicting psychopathology. Previous results suggest that carriers of the TT genotype of rs1360780 who were exposed to child abuse show differences in structure and functional activation of emotion-processing brain areas belonging to the salience network. Extending these findings on intermediate phenotypes of psychopathology, we examined if the interaction between rs1360780 and child abuse predicts resting-state functional connectivity (rsFC) between the amygdala and other areas of the salience network. We analyzed data of young European adults from the general population (*N* = 774; mean age = 18.76 years) who took part in the IMAGEN study. In the absence of main effects of genotype and abuse, a significant interaction effect was observed for rsFC between the right centromedial amygdala and right posterior insula (*p* < .025, FWE-corrected), which was driven by stronger rsFC in TT allele carriers with a history of abuse. Our results suggest that the TT genotype of rs1360780 may render individuals with a history of abuse more vulnerable to functional changes in communication between brain areas processing emotions and bodily sensations, which could underlie or increase risk for psychopathology.

## Introduction

Several lines of research suggest that genetic predisposition as well as child abuse are important risk factors for psychopathology (e.g., McCrory et al., 2012), and that the interaction of these factors can account for variance above main effects of genotype and environment (Belsky & Pluess, 2009; Rutter et al., 2006). In a recent meta-analysis spanning 14 studies with over 15,000 participants, Wang and colleagues (2018) found strong evidence of interactions between *FKBP5* genotypes and early-life stress contributing to the risk of major depressive disorder (MDD) and post-traumatic stress disorder (PTSD). In order to clarify mechanisms by which gene-environment interaction (GxE) leads to psychopathology, attention has turned to intermediate phenotypes - quantitative biological traits that are heritable and associated with a clinical phenotype (Rasetti & Weinberger, 2011). While regional alterations in brain structure and functional activation have commonly been studied as intermediate phenotypes, less attention has been drawn to alterations in brain connectivity. The present study therefore aims to investigate the interactive effects of child abuse and FKBP5 genotype on amygdala resting-state functional connectivity (rsFC), a potential intermediate phenotype for (vulnerability to) psychopathology (e.g., Mulders et al., 2015).

One of the abundantly described psychobiological consequences of child abuse is dysregulation of the hypothalamic-pituitary-adrenal (HPA) axis (McCrory et al., 2010, 2012). The HPA axis is a neuroendocrine system that is involved in the stress response. Exposure to stress triggers the release of glucocorticoid hormones such as cortisol, which binds to mineralocorticoid (MR) and glucocorticoid (GR) receptors. Cortisol preferentially binds to MRs because of their high affinity, even when low levels of cortisol are present. In contrast, GRs, which have about one tenth the affinity of MRs, are occupied mostly when high levels of cortisol are present, e.g., during acute stress responses (Reul & de Kloet, 1985). GRs also play a role in terminating the stress response via a negative feedback loop. The binding of cortisol to GRs at different levels of the HPA axis suppresses activity of the axis and allows cortisol to return to baseline levels (de Kloet, 1991; Tsigos & Chrousos, 2002).

A proper functioning of the negative feedback loop of the HPA axis seems to be critical for a healthy stress response by protecting the body from potential damage caused by the toxic effects of stress hormones. In contrast, dysfunction of the stress response is considered a core characteristic of several stress-related psychiatric disorders. A meta-analysis revealed that patients with MDD had much higher cortisol levels during the recovery period from a stressor than their non-depressed counterparts (Burke et al., 2005). This is thought to be related to an impairment in GR-mediated negative feedback which could result from a decreased GR-sensitivity to glucocorticoids (Pariante, 2004).

It is now well established that genetic factors influence the GR-mediated regulation of the stress response. A central role is attributed to the FKBP5 gene, which is located on chromosome 6p21, and codes for FK506 binding protein 51 (FKBP5), a co-chaperone of the heat shock protein (hsp) 90 that regulates GR sensitivity. FKBP5 is mainly expressed in the brain, including regions involved in the stress response such as the amygdala (Scharf et al., 2011). When FKBP5 is bound to the GR-complex via hsp90, cortisol binds with lower affinity, and translocation of the GR complex to the nucleus is impeded (Binder, 2009; Denny et al., 2000; Wochnik et al., 2005). Thus, enhanced expression of FKBP5 can impair the negative feedback regulation of the HPA axis, resulting in a prolonged stress response.

Increased lymphocyte FKBP5 protein levels have been observed in TT compared to CT/CC allele carriers of rs1360780, a common single nucleotide polymorphism (SNP) within the FKBP5 gene (Binder et al., 2004). In line with the suggestion that reduced GR sensitivity would lead to a prolonged stress response, higher cortisol levels during recovery after stress have been observed in healthy TT carriers (Ising et al., 2008), paralleling the observation in individuals with MDD (Burke et al., 2005). Given these findings, dysregulation of the HPA axis, associated with both exposure to child abuse and FKBP5 risk genotype, may be exaggerated by their interaction. Over time, dysregulation of the HPA axis may render TT genotype carriers with a history of child abuse particularly vulnerable for developing changes in brain structure and function that affect behavior, emotion, and learning. In turn, altered functioning of neural circuits could confer risk for stress-related psychopathology (see Matosin et al., 2018).

One of the core dysfunctions in psychopathology is altered emotion processing. In MDD and PTSD, this commonly includes an attentional bias toward and increased neural activity in response to negative emotional cues (Etkin & Wager, 2007; Hayes et al., 2012; Leppänen, 2006; Shin et al., 2005). Given the importance of the amygdala in emotion processing (Phelps & LeDoux, 2005), interaction studies of FKBP5 and early-life stress have focused on alterations in amygdala activity and volume as promising intermediate phenotypes of psychopathology. In a study on adolescents, White et al. (2012) demonstrated that rs1360780 interacts with childhood adversity to predict threat-related amygdala reactivity, assessed during an emotional face-matching task. Reactivity of the dorsal amygdala increased with the level of retrospectively reported emotional neglect in TT/CT, but not in CC carriers. This is in accordance with a recent study by Holz and colleagues (2015), who used the same task in a high-risk community sample of healthy young male adults and found an interaction effect of FKBP5 with emotional neglect on amygdala reactivity in the same direction as White et al. (2012). Further, the authors reported that amygdala-hippocampus connectivity assessed during the threat-reactivity task decreased with emotional neglect in CC carriers and increased in TT carriers, with CT carriers being intermediate. However, this association was only significant at a liberal threshold and did not survive small volume correction. Although not many studies are available yet, this preliminary evidence supports the hypothesis that early stressful experiences increase the likelihood for disturbed emotion processing, particularly in carriers of the risk allele of rs1360780.

More recently, Grabe and colleagues (2016) reported structural brain differences in adults with a history of abuse who carry the TT genotype of rs1360780 in subcortical and cortical emotion-processing areas, including reduced grey matter volumes in the amygdala, hippocampus, anterior and middle cingulate cortex, and insula. These areas correspond to a large extent to the salience network (SN), which has been linked to processing of various salient stimuli, including stress-related ones (Hermans et al., 2014), and assessment of stimulus relevance in order to guide behavior (Seeley et al., 2007). Specifically, the SN includes the amygdala, fronto-insular cortex, dorsal anterior cingulate cortex (ACC) and temporal poles (Seeley et al., 2007).

The amygdala responds to environmental challenges such as threat, and signals this information to the brainstem, thereby initiating neuroendocrine and autonomic responses (Davis & Whalen, 2001; LeDoux, 2007). The dorsal ACC is involved in cognitive control by modulating amygdala activity, as during reappraisal of negative emotion (Ochsner & Gross, 2005). Together with the ACC, the insula gives rise to feelings and motivations underlying emotions (Medford & Critchley, 2010). Given these functions of areas involved in the SN, it has been suggested that dysfunction within the SN can account for impairments in emotion processing, cognition, autonomic regulation, and neuroendocrine responses, which may underlie heightened emotional reactivity in psychopathology (Price & Drevets, 2010).

Previous research has shown that communication between brain regions ascribed to the SN is altered in individuals with a history of child maltreatment. Specifically, reduced coupling of the amygdala with the medial-orbital prefrontal cortex, cingulate cortex, hippocampus and insula has been found in maltreated individuals in resting state (for review, see Teicher et al., 2016). However, maltreatment-related findings on rsFC are quite heterogeneous, which might be related to the fact that studies investigated different subtypes of maltreatment (i.e., abuse vs. neglect vs. combined), and chose different seed regions in connectivity analyses. Regardless of the direction of reported connectivity differences, studies do suggest that child maltreatment leads to a dysfunctional communication between brain regions when the individual is at rest, which could confer vulnerability for psychopathology.

Taken together, the literature demonstrates that alterations in rsFC in the SN occur in individuals with a history of maltreatment (Teicher et al., 2016). At the same time, such alterations may also represent a core dysfunction in psychiatric disorders such as MDD (Mulders et al., 2015) and PTSD (Koch et al., 2016). Further, it has been suggested that child maltreatment increases risk for psychopathology in carriers of the risk genotype of rs1360780 particularly, likely through its impact on sensitization of the stress response and consequently development of altered brain structure and function (Binder et al., 2009; Matosin et al., 2018).

So far, no study has investigated rsFC of the amygdala in the context of early-life stress by considering FKBP5 rs1360780 genotype. Given that Grabe et al. (2016) found structural differences in brain regions of the SN in TT carriers of rs1360780 with a history of child abuse, the present study aims to extend these findings by examining rsFC between subnuclei of the amygdala and other brain regions of the SN, specifically the ACC and the insula. In order to take into account the structural and functional heterogeneity of the amygdala (Phelps & LeDoux, 2005), the basolateral amygdala (BLA) and the centromedial amygdala (CMA) were chosen as seed regions for connectivity analyses. While the BLA affectively evaluates sensory information (Jovanovic & Ressler, 2010), the CMA is involved in the fear response (LeDoux, 1998). In line with the finding of Teicher et al. (2016), we hypothesized that individuals with a history of child abuse carrying the TT allele of rs1360780 would show altered coupling between seed regions of the amygdala, specifically the CMA, and regions of the SN, including the insula and the ACC, compared with CT/CC allele carriers with and without a history of child abuse.

## Materials and Methods

### Participants

We analyzed data from the longitudinal IMAGEN study (Schumann et al., 2010) comprising European adolescents from Germany, the United Kingdom, Ireland, and France. At baseline, 2,462 individuals participated at the age of 14 and, to date, have been followed up three times. Data were collected from eight study sites (Berlin, Dresden, Dublin, Hamburg, London, Mannheim, Nottingham, and Paris). We included all participants from IMAGEN’s second follow-up wave, from whom resting-state data, childhood adversity status, and genotype were available (*N* = 819). Data from the second follow-up were used specifically, because childhood adversity, as measured by the Childhood Trauma Questionnaire, was not measured at baseline and more resting-state data sets were available for the second follow-up. Thirty-two participants had to be excluded due to insufficient MRI data quality, as identified by visual inspection, and 13 due to extensive movement during scanning (>3 mm of translation; >1° of rotation). The final sample thus comprised 774 participants. The sample characteristics of the final sample are shown in Table 1.

**Table 1.** Descriptive characteristics for the IMAGEN sample

| Characteristics | Total sample | Group |  |  | Test statistics for comparison of TT vs CT/CC <sup>a</sup> | df<sup>a</sup> | p-value <sup>a</sup> |
| --- | --- | --- | --- | --- | --- | --- | --- |
|  |  | rs1360785 = CC | rs1360785 = CT | rs1360785 = TT |  |  |  |
| *N* | 774 | 382 | 319 | 73 |  |  |  |
| **Age (range)** | 18.76 (17-23) |  |  |  |  |  |  |
| **Sex**<br>(male/female) | 378/396 | 190/192 | 153/166 | 35/38 | $\chi^2 = .026$ | 1 | .873 |
| **Handedness**<br>(right/left) | 693/81 | 336/46 | 291/28 | 66/7 | $\chi^2 = .066$ | 1 | .797 |
| **Childhood abuse**<br>(yes/no) <sup>b</sup> | 504/270 | 250/132 | 206/113 | 48/25 | $\chi^2 = .014$ | 1 | .904 |
| **CTQ total score**<br>(*M* ± *SD*) | 31.55±7.07 | 32.04±7.50 | 30.92±5.80 | 31.78±9.37 | *t* = -.830 | 772 | .774 |
| **CTQ abuse score**<br>(*M* ± *SD*) | 17.60±3.98 | 17.80±4.34 | 17.29±3.14 | 17.97±5.10 | *t* = -.287 | 772 | .407 |

Annotations: *N* = sample size; *M* = mean; *SD* = standard deviation; <sup>a</sup> According to two-sample t-test (two-tailed) for continuous variables or $\chi^2$ tests for categorical variables to check for possible differences in the genotype groups (CC/CT vs. TT).; <sup>b</sup> Childhood abuse derived from the Childhood Trauma Questionnaire (none versus any).

The Development and Well-Being Assessment (DAWBA) was used to assess psychopathology (Goodman et al., 2011). In short, computer-predicted diagnoses were generated for a range of disorders (i.e., 50% or more chance of having the actual disorder in reference to a norm group). An indication for a diagnosis was given for 115 out of the 774 young adults, with 31 individuals having had an indication for more than one diagnosis. Depression and anxiety disorders were most prevalent (depression *n* = 60, generalized anxiety disorder *n* = 23, social phobia *n* = 23, eating disorder *n* = 15, panic disorder *n* = 12, specific phobia *n* = 9, posttraumatic stress disorder *n* = 5, tic disorder *n* = 5, agoraphobia *n* = 2, conduct disorder *n* = 1, and obsessive compulsive disorder *n* = 1). While the vast majority of the indications fell into the 50-70% range of having the actual disorder, 22 indications fell in the 70% or higher range for depression specifically.

The study was performed in accordance with the Declaration of Helsinki and approved by local ethics committees at each site. Written informed consent was obtained from all participants. Participants received financial compensation for their participation and travel expenses. A precise description of recruitment and assessment procedures spanning exclusion and inclusion criteria has been published elsewhere (Schumann et al., 2010).

### Assessment of child abuse

The Childhood Trauma Questionnaire (CTQ) was used to assess childhood adversity. The CTQ has good reliability and validity, as was demonstrated in independent studies (Bernstein et al., 2003; Wingenfeld et al., 2010). A total of 28 items is rated on a five-point Likert scale, ranging from 1 (never true) to 5 (very often true), with higher scores indicating a higher exposure to traumatic experiences during childhood. The three abuse subscales (i.e., emotional, sexual, and physical abuse) were used to calculate an abuse sum score. Following the approach of Grabe et al. (2016), based on the CTQ scoring manual, we generated a dichotomized variable of overall abuse by which a participant was rated as positive for abuse when any of the abuse subscales had a sum score indicative of at least moderate abuse (i.e., ≥ 9 for emotional abuse, ≥ 8 for physical abuse, and ≥ 6 for sexual abuse). However, as according to this classification only 18 T-homozygotes in our sample could be categorized as having experienced child abuse, we decided to apply a more lenient threshold than Grabe et al. (2016). Specifically, a participant was categorized as positive for overall abuse when a score of at least two was reported on either one of the abuse items, therefore separating participants without a history of abuse from the ones who were exposed to any type and intensity of abuse. While this increased the sample size of T-homozygotes with a history of child abuse considerably, the inclusion of these very mild cases also rendered us less sensitive to find effects of abuse.

### Genotyping

A precise description of genotyping and imputation procedures has been published elsewhere (Lett et al., 2020; Schumann et al., 2010). In brief, blood samples were collected during site visits and sent to the IMAGEN DNA biobank for processing. Analyses covered DNA, for which the Illumina Quad 610 chip and 660w chip (Illumina, San Diego, CA, USA) were used to perform genome-wide genotyping of about 600mult.000 SNPs. Genotype results for the FKBP5 gene SNP rs1360780 were derived from standard imputation procedures. The distribution of rs1360780 alleles did not deviate from the Hardy-Weinberg equilibrium (χ2 = 0.29, *p* = .59). As per Grabe et al. (2016) and other studies on rs1360780 genotype (Koopmann et al., 2016), we combined the CC and CT carriers into one group, thus comparing T-homozygotes with C allele carriers.

### MRI data acquisition

Structural and functional MRI data were acquired on 3T whole body MR scanners from different manufacturers (Siemens, Munich, Germany; Philips, Best, The Netherlands; GE Healthcare, Chicago, USA; Bruker, Ettlingen, Germany). For each sequence, parameters directly affecting image contrast or signal-to-noise ratio were determined and held constant across sites to minimize differences between scanners (Schumann et al., 2010). Functional MRI measurements were performed using a single-shot T2*-weighted gradient-echo echoplanar imaging (GE-EPI) sequence with 3.4 x 3.4 mm in-plane voxel size, 164 volumes, 40 slices with a thickness of 2.4 mm with a gap of 1.0 mm (3.4 mm total intra-slice distance), repetition time of 2200 ms, echo time of 30 ms, a flip angle of 75°, and a Field of View of 218 x 218 mm. Volumes were acquired in sequential ascending slice order. For the resting-state scan participants were instructed to lie still with their eyes closed and let their minds wander without focusing on a specific thought. A standard T1-weighted structural volume with sagittal volume excitation and 1.1 x 1.1 x 1.1 mm^3^ voxel size was created for every participant for subsequent normalization purposes. All T1-weighted images were screened for incidental, clinically relevant findings by a neuroradiologist prior to analysis.

### fMRI data preprocessing

First, all imaging data were visually screened for corrupted data or acquisition artifacts. FEAT (fMRI Expert Analysis Tool) Version 6.00 in FSL (FMRIB Software Library v5.0, www.fmrib.ox.ac.uk/fsl) was used to perform data preprocessing on the functional data, including motion correction (Jenkinson et al., 2002), slice time correction, non-brain removal (Smith, 2002), spatial smoothing using a Gaussian kernel of 4 mm FWHM (full width of half maximum), and grand mean intensity normalization. Independent Component Analysis (ICA)-based automatic removal of motion-related and physiological noise artifacts was used to further clean the data (ICA-AROMA; Pruim et al., 2015). Next, data were high-pass temporal filtered (> 0.008 Hz) to remove slow drifts. The middle EPI volume was co-registered to the individual brain-extracted T1 image, using boundary-based registration (Greve & Fischl, 2009). Non-linear normalization of the T1 image to the 2mm MNI standard space template (Montreal Neurological Institute, Quebec, Canada) was done using Advanced Normalization Tools (ANTs; Avants et al., 2008). Last, the fully preprocessed data were normalized to 2 mm MNI standard space, applying the registration matrices and warp images from the two previous registration steps.

### fMRI data analysis

To study amygdala functional connectivity, a seed-based connectivity analysis was carried out. Binary seed masks of the CMA and BLA nucleus of the left and right amygdala were created, which only included voxels with a probability higher than 50% using the Juelich Histological Atlas, as provided in FSLview. For each of the four masks, the first Eigen time series was obtained from the preprocessed resting-state data. To generate functional connectivity maps, the time series of each participant’s right and left CMA and BLA were regressed separately against every other voxel’s time series using the general linear model (GLM) with FSL’s command line tool fsl_glm. Time series extracted from the CSF and deep white matter were included in the model as nuisance variables.

The individual whole-brain connectivity maps were fed into a higher-level GLM, for the left and right CMA and BLA separately, using the dichotomized CTQ abuse variable, FKBP5 genotype, and their interaction as regressors of interest, adding age (continuous) and scan site, sex, and DAWBA diagnosis (dummy coded) covariates. The resulting *t*-statistical maps subsequently underwent Threshold-Free Cluster Enhancement (TFCE) (Smith & Nichols, 2009), which describe the association between connectivity and CTQ score, using the default parameter settings (H = 2, E = 0.5, C = 6). Significance testing was carried out with permutation testing (10,000 iterations) using the software suite *TFCE_mediation* (https://github.com/trislett/tfce_mediation; Lett et al., 2017). In this step, the true findings were tested against a null distribution of random results, which was generated beforehand. This resulted in statistical images that are family-wise error corrected for multiple comparisons across all voxels and Bonferroni-corrected for assessing two subnuclei at *p* < .025. Given our a priori expectations of differences in areas of the SN, we created a ROI mask of the bilateral insula and ACC, using the Harvard-Oxford Cortical Structural Probability Atlas, as provided in FSLview. To be as unbiased as possible, no probability threshold was used for these regions, and corrections for multiple comparisons were therefore done on all voxels that had any probability of being part of these regions. Given the effects on grey matter volume observed by Grabe et al.

(2016), we also controlled for grey matter volume as it could potentially drive connectivity effects. To this end, FSL’s command line tool feat_gm_prepare was used to produce a voxelwise confound regressor: First, structural scans were grey matter segmented using FAST (fMRI’s Automated Segmentation Tool; Zhang et al., 2001); second, the previously generated warp files were applied to the grey matter maps, concatenated across participants, and demeaned.

The amygdala seeds and ROI mask, as well as the corrected and uncorrected statistical images of our analyses can be found under: https://neurovault.org/collections/7224.

## Results

The final sample of 774 participants comprised 245 CC/CT allele carriers without a history of abuse, 456 CC/CT allele carriers with a history of abuse, 25 TT allele carriers without a history of abuse, and 48 TT allele carriers with a history of abuse. Sample characteristics are reported in Table 1. Importantly, the dichotomized criterion for abuse (i.e., none vs. any) was equally distributed across genotypes (CC/CT vs. TT; *p* = .904). A history of abuse was more frequently observed in young adults with than without an indication of psychopathology, *χ*^*2*^(1, *N* = 774) = 10.782, *p* = .001.

Average functional connectivity maps of the CMA and BLA across all participants are shown in Supplemental Figure 1. We neither found a main effect of genotype or child abuse on amygdala rsFC in either the whole-brain or ROI analyses (*p*_FWE_ > .05 for all comparisons), nor an interaction between genotype and child abuse in the whole-brain analyses for any of the seeds (*p*_FWE_ > .025 for all comparisons). However, a significant interaction was found between child abuse and FKBP5 genotype on rsFC of the right CMA with the right posterior insula (x = 36; y = −12; z = −2), corrected for multiple comparisons (*p* < .025; see Figure 1A). The interaction effect was driven by stronger amygdala rsFC with the insula in TT allele carriers with a history of abuse. Post-hoc *t*-tests demonstrated that CMA rsFC with the posterior insula significantly differed between CC/CT allele carriers without a history of abuse and TT allele carriers without a history of abuse, *t*(32.024)= 2.73, *p* = .01, and between TT allele carriers with and without a history of abuse, *t*(71) = −2.88, *p* = .005 (see Figure 1B). Importantly, inclusion of grey matter volume or DAWBA diagnosis as covariate did not affect the results, as the same region was found significant without adjustment for these factors as well.

**Figure 1.**
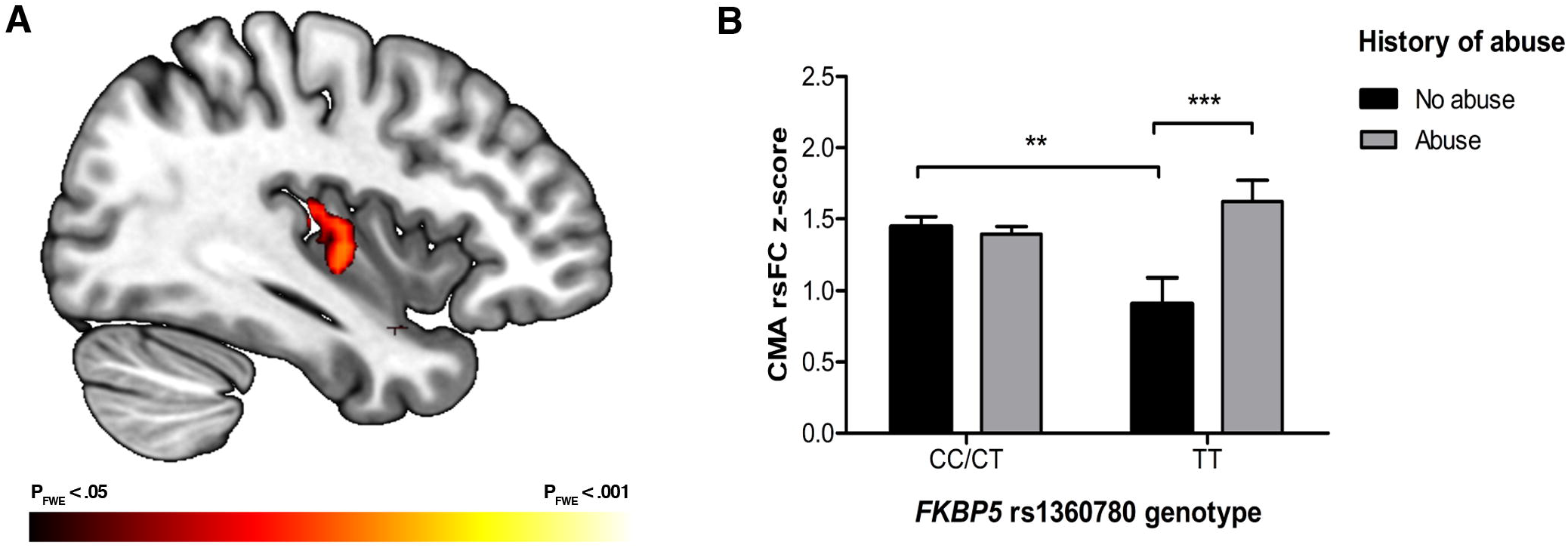
**(A)** Interaction effect of FKBP5 and child abuse on centromedial amygdala resting-state functional connectivity with the right posterior insula overlaid on the MNI standard space template (at x = 38), shown at a significance threshold of *p*_FWE_ < .05 for illustration purposes. A more restricted part of this cluster was found when applying the stringent correction for both the number of voxels and number of amygdala subregions (*p*_FWE_ < .025). **(B)** Bar graph illustrating the significant interaction effect of FKBP5 rs1360780 with child abuse on resting-state functional connectivity between the right centromedial amygdala (CMA) and right posterior insula. Error bars represent the standard error of the mean (SEM). *p*-value for two-sample *t*-tests between indicated groups ** = .01; *** =.005.

## Discussion

In the present study, we investigated the impact of child abuse, genetic variation within the FKBP5 gene, and their interaction on functional connectivity within the salience network during resting state. A significant interaction effect emerged on rsFC between the right CMA and the right posterior insula. Specifically, carriers of the TT genotype of rs1360780 with a history of child abuse demonstrated a stronger coupling between these regions compared to carriers of the TT genotype of rs1360780 without a history of child abuse. Further, in individuals without a history of child abuse, TT allele carriers showed a weaker coupling between these regions compared to CC/CT allele carriers. No main effect of either genotype or child abuse was present. Contrary to our expectations, no effects were found for rsFC within the dACC, the other core region of the salience network.

Our results add important evidence to GxE influences on intermediate phenotypes of psychopathology by demonstrating that alterations in rsFC occur in genetically vulnerable individuals with experiences of abuse. We observed a stronger rsFC between the posterior insula and the amygdala—brain regions that contribute to bodily sensations and arousal. The posterior insula, primarily connected to sensorimotor, posterior temporal, and parietal areas, has been ascribed the role of sensorimotor integration and is seen as critical for interoception (Cauda et al., 2011; Craig, 2002; Deen et al., 2011; Kurth et al., 2010; Uddin et al., 2017). More specifically, Craig (2009) proposed that the posterior insula hosts a representation of primary interoceptive information, which is re-represented and integrated with affective aspects in a polymodal zone situated in mid and anterior parts of the insula. Following a posterior-to-mid-to-anterior pattern, the successive integration of information stemming from various modalities, including emotionally salient environmental stimuli, hedonic, motivational, and social conditions, is posited to underlie the current feeling and awareness of oneself (Craig, 2009). The CMA is critically involved in the neural stress response, as it affects the release of cortisol and modulates the functioning of the autonomic nervous system through its connections to the hypothalamus (Davis, 1992; Jovanovic & Ressler, 2010; LeDoux, 1998). Given that a stronger connectivity indicates greater signal covariance between regions of interest, activation of the CMA and the posterior insula may be more coordinated in genetically vulnerable individuals with experiences of abuse, even in the absence of a stressful situation. Based on our results, we thus speculate that the amygdala may alter posterior insula representation of primary interoceptive information, leading to increased processing of bodily feelings with respect to emotions in TT allele carriers with a history of abuse.

According to contemporary neurobiological models (Menon, 2011), aberrant interactions within and between neurocognitive networks may underlie or predispose individuals to disrupted cognitive processes, which in turn can lead to psychopathology. Disruptions in amygdala-insula connectivity have been reported in the context of generalized anxiety disorder (Roy et al., 2013), PTSD (Nicholson et al., 2016; Rabinak et al., 2011; Sripada et al., 2012), and MDD (Jacobs et al., 2016; Veer et al., 2010). Given the largely non-clinical sample used in our study, while controlling for DAWBA diagnosis indication in the analyses, the alterations in rsFC may represent a precursor of clinically relevant symptoms, mediating enhanced vulnerability to stress-related psychopathology. However, although TT allele carriers with a history of abuse did demonstrate the strongest connectivity levels, these were statistically not significantly different from connectivity levels of CC/CT allele carriers with and without a history of abuse. Future follow-up analyses of the present sample should shed more light on whether stronger amygdala-insula rsFC can indeed be considered a vulnerability marker.

While the interaction effect observed in our study emerged between the amygdala and posterior insula, it is interesting to note that former connectivity studies investigating correlates of trauma (Thomason et al., 2015) or psychopathology (Jacobs et al., 2016; Veer et al., 2010) have found alterations in the coupling between the amygdala and the anterior insula. Although it has been shown that the anterior and posterior insula belong to complementary networks with different connectivity patterns and functions (Cauda et al., 2011; Deen et al., 2011), little agreement exists regarding boundaries or functional specialization of insula subregions across studies (Gasquoine, 2014; Kurth et al., 2010). Given that the anterior and posterior insula are thought to interact to modulate autonomic reactivity to salient stimuli (Menon, 2011; Menon & Uddin, 2010), the interaction effect observed in our study could also reflect an exaggerated state of arousal despite the absence of an actual danger in TT allele carriers with experiences of abuse. Given that several studies (e.g., Rabinak et al., 2011; Roy et al., 2013) have not separated insula subregions, the role of anterior and posterior insula connectivity to the amygdala in the context of psychopathology needs further investigation.

It is interesting to note that in our study, TT allele carriers without a history of abuse exhibited the lowest amygdala-insula rsFC. The interaction effect thus resembles a differential-susceptibility pattern (Belsky & Pluess, 2009), in which the risk genotype could function as a “plasticity factor”. One could speculate that the TT allele renders an individual more vulnerable than others to the negative effects of adversity, leading to increased amygdala-insula rsFC. On the other hand, it may also render an individual disproportionately susceptible to the beneficial effects of a non-adverse environment, as indicated by weaker connectivity strength. Previously, it has been proposed that FKBP5 alleles may follow the differential susceptibility theory (Matosin et al., 2018), although findings differ with regards to whether the TT genotype or CT/TT genotypes serve as plasticity factors. A recent GxE study on children with early institutional care has shown that girls with the CT/TT genotype exhibited more depressive symptoms at higher levels of peer victimization, but fewer depressive symptoms at lower levels of peer victimization compared to girls carrying the CC allele (VanZomeren-Dohm et al., 2015). In addition, the study of Binder et al. (2004) demonstrated that despite experiencing higher rates of lifetime depressive episodes, TT allele carriers are more responsive to treatment with antidepressant medication compared to CC/CT allele carriers. Given that the functional significance of low amygdala-insula connectivity levels during resting state remains unclear, we cannot infer whether our findings point towards beneficial effects of the TT allele. However, we speculate that a stronger connectivity of these regions may be detrimental, as previous studies have revealed similar alterations in the context of PTSD (Rabinak et al., 2011; Sripada et al., 2012).

As Grabe and colleagues (2016) reported on reduced grey matter volumes in the ACC in TT allele carriers with a history of child of abuse, we also expected to observe GxE-related alterations in functional coupling between the amygdala and ACC. Importantly, such effects have previously been observed in the context of stress-related disorders (Anand et al., 2005; Brown et al., 2014; Carballedo et al., 2011; Pannekoek et al., 2014), which were suggested to underlie impairments in salience processing and emotional modulation in these disorders. However, we did not find a GxE relation with amygdala-ACC connectivity in our sample, which might be due to the largely non-clinical nature of our sample with predominantly low experiences of abuse.

Although childhood maltreatment has repeatedly been associated with alterations in functional connectivity (Dean et al., 2014; Fonzo et al., 2013; Jedd et al., 2015; Teicher et al., 2016; van der Werff et al., 2013), we did not find a main effect of abuse in our study. Also genotype itself was not related with connectivity outcomes. Our finding of an interaction effect in absence of main effects supports the notion that the interplay of genetic vulnerability and stressful experiences, even at a low severity level, may affect the communication between brain networks at rest. This is in line with literature demonstrating that only few studies have reported main effects of FKBP5 on risk for psychopathology, whereas stronger evidence points towards interaction effects with early life stress (Matosin et al., 2018; Wang et al., 2018).

Recent research (Klengel et al., 2013; Klengel & Binder, 2015; Matosin et al., 2018) describes molecular mechanisms that explain how the interaction of FKBP5 and childhood adversity could shape intermediate phenotypes of psychopathology: In the context of GR stimulation, the T allele facilitates FKBP5 expression, which impedes cortisol-GR binding and consequently impairs the negative feedback loop of the HPA axis. This effect is likely exacerbated by T allele-specific stress-related demethylation at regulatory glucocorticoid responsive elements, further enhancing FKBP5 expression in the context of GR activation and intensifying impairments in HPA axis functioning among T allele carriers. In line with this model (Matosin et al., 2018), associations between circulating levels of cortisol and amygdala activation and functional connectivity have been documented (Bogdan & Hariri, 2012; Burghy et al., 2012; Urry, 2006; Veer et al., 2012), supporting the hypothesis that alterations in brain connectivity observed in the TT allele carriers in our study may reflect long-term consequences of a functional change in the HPA axis.

The present study contributes to a burgeoning literature demonstrating interaction effects of FKBP5 with early adversity on intermediate phenotypes of psychopathology: Grabe and colleagues (2016) reported on widespread structural brain differences in TT carriers, including reduced grey matter volumes of the amygdala and insula. Further, two studies found that CT/TT allele carriers showed increasing threat-related reactivity in the right amygdala with the level of emotional neglect (Holz et al., 2015; White et al., 2012). Recently, interaction effects of FKBP5 with non-adverse environmental conditions were demonstrated by Matsudaira and colleagues (2019). Based on a sample of Japanese children, they showed that TT carriers had a reduced thalamic gray matter volume compared with CC carriers at low to moderate levels of maternal acceptance. Although accumulating evidence points towards increased risk for structural and functional brain alterations in the context of FKBP5 interaction with environmental conditions, longitudinal studies are needed to close the gap between risk factors, intermediate phenotypes, and occurring clinical symptoms.

## Limitations

There are several limitations of our study that need to be considered when interpreting the results. First, the number of individuals reported having experienced severe abuse was relatively small, which rendered us unable to investigate abuse-specific interaction effects with FKBP5. By applying a more lenient threshold to group classification of individuals, we instead focused on any type of abuse, including “minimal” experiences of abuse. A further study following children at risk is needed to examine whether the observed heightened amygdala-insula rsFC occurs specifically in genetically vulnerable individuals with more severe experiences of abuse. Given a frequency of approximately 10% of FKBP5 TT-homozygotes, such a study would warrant inclusion of a large number of participants.

Second, no objective measure of child abuse was included in our study. However, this limitation may not be as problematic as generally expected (for discussion, see Teicher et al., 2016). Severe cases of abuse have been shown to be most likely identified by both prospective case reviews and retrospective self-reports, whereas less severe incidents are more likely to be missed by both (Shaffer et al., 2008). If this holds true, the interaction effect observed in our study might even be an underestimation. Further, claims have been made that retrospective self-reports as opposed to official records of maltreatment show unique relations to physical and mental health outcomes (Kendall-Tackett & Becker-Blease, 2004). Future investigations could clarify whether different intermediate phenotypes arise when interactions between risk genotype with prospective versus retrospective measures of abuse are investigated.

Third, only one polymorphism within the FKBP5 gene was investigated. The SNP rs1360780 was chosen based on our intention to extend findings of Grabe and colleagues (2016). Meta-analytic evidence further supports the important role of rs1360780 interacting with early life stress in increasing risk for depression or PTSD (Wang et al., 2018). However, this does not exclude the possibility that different polymorphisms within the FKBP5 gene or polymorphisms from other genes could interact with childhood abuse on rsFC as well (see Hart et al., 2017; Pagliaccio et al., 2015).

Fourth, no endocrine data such as cortisol was collected from our participants, which could have shed light on possible HPA axis alterations in genetically vulnerable individuals with experiences of child abuse, thereby allowing insights in the relation between central and peripheral effects of FKBP5.

Last, interaction effects between FKBP5 and adversity observed in our study relate to resting state, and thus cannot be used to infer network abnormalities during the processing of specific stimuli or performance of a specific task. For this purpose, both resting state and task-dependent neuroimaging should be implemented which could provide differential insights in brain functioning in the context of adversity.

## Conclusion

In summary, the present study demonstrates interaction effects of FKBP5 rs1360780 with child abuse on functional connectivity during resting state. Only in TT allele carriers, the experience of abuse, although mild, was associated with a stronger coupling between the amygdala and posterior insula, which could reflect an enhanced processing of bodily feelings with respect to emotions in the absence of danger. Such abuse-related functional connectivity patterns may constitute intermediate phenotypes of psychopathology and could provide a useful target for interventions that promote psychological well-being after adverse experiences.

## Data Availability

The data that support the findings of this study are available on request from the corresponding author. The data are not publicly available due to privacy or ethical restrictions.

## Authors’ contribution statement

TB, GJB, ALWB, EBQ, SD, HF, AG, HG, RB, JLM, EA, FN, DPO, LP, SH, JHF, MNS, RW, GS, AH, HW have made substantial contributions to conception and design, or acquisition of data. CW, IMV, NYL, LSD, TAL, HW have made substantial contributions to the analysis and interpretation of data and were involved in drafting the manuscript. All authors approved to be included in the author list, revised the manuscript critically for important intellectual content, and gave written consent for publication in its current form.

## Acknowledgments

CW is supported by the UvA-FMG Research Priority Area (RPA) Yield. IMV and HW are supported by the German Research Foundation (Deutsche Forschungsgemeinschaft), grant number WA1539/7-1. This work furthermore received support from the following sources: the European Union-funded FP6 Integrated Project IMAGEN (Reinforcement-related behaviour in normal brain function and psychopathology) (LSHM-CT-2007-037286), the Horizon 2020 funded ERC Advanced Grant ‘STRATIFY’ (Brain network based stratification of reinforcement-related disorders) (695313), Human Brain Project (HBP SGA 2, 785907, and HBP SGA 3, 945539), the Medical Research Council Grant ‘c-VEDA’ (Consortium on Vulnerability to Externalizing Disorders and Addictions) (MR/N000390/1), the National Institute of Health (NIH) (R01DA049238, A decentralized macro and micro gene-by-environment interaction analysis of substance use behavior and its brain biomarkers), the National Institute for Health Research (NIHR) Biomedical Research Centre at South London and Maudsley NHS Foundation Trust and King’s College London, the Bundesministeriumfür Bildung und Forschung (BMBF grants 01GS08152; 01EV0711; Forschungsnetz AERIAL 01EE1406A, 01EE1406B; Forschungsnetz IMAC-Mind 01GL1745B), the Deutsche Forschungsgemeinschaft (DFG grants SM 80/7-2, SFB 940, TRR 265, NE 1383/14-1), the Medical Research Foundation and Medical Research Council (grants MR/R00465X/1 and MR/S020306/1), the National Institutes of Health (NIH) funded ENIGMA (grants 5U54EB020403-05 and 1R56AG058854-01). Further support was provided by grants from: – the ANR (ANR-12-SAMA-0004, AAPG2019 – GeBra), the Eranet Neuron (AF12-NEUR0008-01 – WM2NA; and ANR-18-NEUR00002-01 – ADORe), the Fondation de France (00081242), the Fondation pour la Recherche Médicale (DPA20140629802), the Mission Interministérielle de Lutte-contre-les-Drogues-et-les-Conduites-Addictives (MILDECA), the Assistance-Publique-Hôpitaux-de-Paris and INSERM (interface grant), Paris Sud University IDEX 2012, the Fondation de l’Avenir (grant AP-RM-17-013), the Fédération pour la Recherche sur le Cerveau; the National Institutes of Health, Science Foundation Ireland (16/ERCD/3797), U.S.A. (Axon, Testosterone and Mental Health during Adolescence; RO1 MH085772-01A1), and by NIH Consortium grant U54 EB020403, supported by a cross-NIH alliance that funds Big Data to Knowledge Centres of Excellence.

## Figures

**Supplemental Figure 1** Average seed-based functional connectivity for the centromedial and basolateral amygdala across all participants (uncorrected, but arbitrarily thresholded at *t* > 15). overlaid on the MNI standard brain. Brains are displayed in radiological convention (i.e., left hemisphere is on the right side of the image and vice versa).

## References

Anand, A., Li, Y., Wang, Y., Wu, J., Gao, S., Bukhari, L., Mathews, V. P., Kalnin, A., & Lowe, M. J. (2005). Activity and connectivity of brain mood regulating circuit in depression: A functional magnetic resonance study. Biological Psychiatry, 57(10), 1079–1088. https://doi.org/10.1016/j.biopsych.2005.02.021

Avants, B. B., Epstein, C. L., Grossman, M., & Gee, J. C. (2008). Symmetric diffeomorphic image registration with cross-correlation: Evaluating automated labeling of elderly and neurodegenerative brain. Medical Image Analysis, 12(1), 26–41. https://doi.org/10.1016/j.media.2007.06.004

Belsky, J., & Pluess, M. (2009). Beyond diathesis stress: Differential susceptibility to environmental influences. Psychological Bulletin, 135(6), 885–908. https://doi.org/10.1037/a0017376

Bernstein, D. P., Stein, J. A., Newcomb, M. D., Walker, E., Pogge, D., Ahluvalia, T., Stokes, J., Handelsman, L., Medrano, M., Desmond, D., & Zule, W. (2003). Development and validation of a brief screening version of the Childhood Trauma Questionnaire. Child Abuse & Neglect, 27(2), 169–190.

Binder, E. B. (2009). The role of FKBP5, a co-chaperone of the glucocorticoid receptor in the pathogenesis and therapy of affective and anxiety disorders. Psychoneuroendocrinology, 34 Suppl 1, S186–195. https://doi.org/10.1016/j.psyneuen.2009.05.021

Binder, E. B., Salyakina, D., Lichtner, P., Wochnik, G. M., Ising, M., Pütz, B., Papiol, S., Seaman, S., Lucae, S., Kohli, M. A., Nickel, T., Künzel, H. E., Fuchs, B., Majer, M., Pfennig, A., Kern, N., Brunner, J., Modell, S., Baghai, T., … Muller-Myhsok, B. (2004). Polymorphisms in FKBP5 are associated with increased recurrence of depressive episodes and rapid response to antidepressant treatment. Nature Genetics, 36(12), 1319–1325. https://doi.org/10.1038/ng1479

Bogdan, R., & Hariri, A. R. (2012). Neural embedding of stress reactivity. Nature Neuroscience, 15(12), 1605–1607. https://doi.org/10.1038/nn.3270

Brown, V. M., LaBar, K. S., Haswell, C. C., Gold, A. L., Mid-Atlantic MIRECC Workgroup, McCarthy, G., & Morey, R. A. (2014). Altered resting-state functional connectivity of basolateral and centromedial amygdala complexes in posttraumatic stress disorder. Neuropsychopharmacology: Official Publication of the American College of Neuropsychopharmacology, 39(2), 351–359. https://doi.org/10.1038/npp.2013.197

Burghy, C. A., Stodola, D. E., Ruttle, p. L., Molloy, E. K., Armstrong, J. M., Oler, J. A., Fox, M. E., Hayes, A. S., Kalin, N. H., Essex, M. J., Davidson, R. J., & Birn, R. M. (2012). Developmental pathways to amygdala-prefrontal function and internalizing symptoms in adolescence. Nature Neuroscience, 15(12), 1736–1741. https://doi.org/10.1038/nn.3257

Burke, H. M., Davis, M. C., Otte, C., & Mohr, D. C. (2005). Depression and cortisol responses to psychological stress: A meta-analysis. Psychoneuroendocrinology, 30(9), 846–856. https://doi.org/10.1016/j.psyneuen.2005.02.010

Carballedo, A., Scheuerecker, J., Meisenzahl, E., Schoepf, V., Bokde, A., Möller, H.-J., Doyle, M., Wiesmann, M., & Frodl, T. (2011). Functional connectivity of emotional processing in depression. Journal of Affective Disorders, 134(1–3), 272–279. https://doi.org/10.1016/j.jad.2011.06.021

Cauda, F., D’Agata, F., Sacco, K., Duca, S., Geminiani, G., & Vercelli, A. (2011). Functional connectivity of the insula in the resting brain. NeuroImage, 55(1), 8–23. https://doi.org/10.1016/j.neuroimage.2010.11.049

Craig, A. D. (2002). How do you feel? Interoception: the sense of the physiological condition of the body. Nature Reviews. Neuroscience, 3(8), 655–666. https://doi.org/10.1038/nrn894

Craig, A. D. (2009). How do you feel--now? The anterior insula and human awareness. Nature Reviews. Neuroscience, 10(1), 59–70. https://doi.org/10.1038/nrn2555

Davis, M. (1992). The Role of the Amygdala in Fear and Anxiety. Annual Review of Neuroscience, 15(1), 353–375. https://doi.org/10.1146/annurev.ne.15.030192.002033

Davis, M., & Whalen, p. J. (2001). The amygdala: Vigilance and emotion. Molecular Psychiatry, 6(1), 13–34.

de Kloet, E. R. (1991). Brain corticosteroid receptor balance and homeostatic control. Front. Neuroendocrinol., 12, 95–164.

Dean, A. C., Kohno, M., Hellemann, G., & London, E. D. (2014). Childhood maltreatment and amygdala connectivity in methamphetamine dependence: A pilot study. Brain and Behavior, 4(6), 867–876. https://doi.org/10.1002/brb3.289

Deen, B., Pitskel, N. B., & Pelphrey, K. A. (2011). Three systems of insular functional connectivity identified with cluster analysis. Cerebral Cortex (New York, N.Y.: 1991), 21(7), 1498–1506. https://doi.org/10.1093/cercor/bhq186

Denny, W. B., Valentine, D. L., Reynolds, p. D., Smith, D. F., & Scammell, J. G. (2000). Squirrel monkey immunophilin FKBP51 is a potent inhibitor of glucocorticoid receptor binding. Endocrinology, 141(11), 4107–4113. https://doi.org/10.1210/endo.141.11.7785

Etkin, A., & Wager, T. D. (2007). Functional neuroimaging of anxiety: A meta-analysis of emotional processing in PTSD, social anxiety disorder, and specific phobia. The American Journal of Psychiatry, 164(10), 1476–1488. https://doi.org/10.1176/appi.ajp.2007.07030504

Fonzo, G. A., Flagan, T. M., Sullivan, S., Allard, C. B., Grimes, E. M., Simmons, A. N., Paulus, M. P., & Stein, M. B. (2013). Neural functional and structural correlates of childhood maltreatment in women with intimate-partner violence-related posttraumatic stress disorder. Psychiatry Research, 211(2), 93–103. https://doi.org/10.1016/j.pscychresns.2012.08.006

Gasquoine, p. G. (2014). Contributions of the insula to cognition and emotion. Neuropsychology Review, 24(2), 77–87. https://doi.org/10.1007/s11065-014-9246-9

Goodman, A., E. Heiervang, S. Collishaw and R. Goodman (2011). The ‘DAWBA bands’ as an ordered-categorical measure of child mental health: Description and validation in British and Norwegian samples. Social Psychiatry and Psychiatric Epidemiology, 46(6), 521–532. https://doi.org/10.1007/s00127-010-0219-x

Grabe, H. J., Wittfeld, K., Van der Auwera, S., Janowitz, D., Hegenscheid, K., Habes, M., Homuth, G., Barnow, S., John, U., Nauck, M., Völzke, H., Meyer zu Schwabedissen, H., Freyberger, H. J., & Hosten, N. (2016). Effect of the interaction between childhood abuse and rs1360780 of the FKBP5 gene on gray matter volume in a general population sample. Human Brain Mapping, 37(4), 1602–1613. https://doi.org/10.1002/hbm.23123

Greve, D. N., & Fischl, B. (2009). Accurate and robust brain image alignment using boundary-based registration. NeuroImage, 48(1), 63–72. https://doi.org/10.1016/j.neuroimage.2009.06.060

Hart, H., Lim, L., Mehta, M. A., Chatzieffraimidou, A., Curtis, C., Xu, X., Breen, G., Simmons, A., Mirza, K., & Rubia, K. (2017). Reduced functional connectivity of fronto-parietal sustained attention networks in severe childhood abuse. PloS One, 12(11), e0188744. https://doi.org/10.1371/journal.pone.0188744

Hayes, J. P., VanElzakker, M. B., & Shin, L. M. (2012). Emotion and cognition interactions in PTSD: A review of neurocognitive and neuroimaging studies. Frontiers in Integrative Neuroscience, 6. https://doi.org/10.3389/fnint.2012.00089

Hermans, E. J., Henckens, M. J. A. G., Joëls, M., & Fernández, G. (2014). Dynamic adaptation of large-scale brain networks in response to acute stressors. Trends in Neurosciences, 37(6), 304–314. https://doi.org/10.1016/j.tins.2014.03.006

Holz, N. E., Buchmann, A. F., Boecker, R., Blomeyer, D., Baumeister, S., Wolf, I., Rietschel, M., Witt, S. H., Plichta, M. M., Meyer-Lindenberg, A., Banaschewski, T., Brandeis, D., & Laucht, M. (2015). Role of FKBP5 in emotion processing: Results on amygdala activity, connectivity and volume. Brain Structure & Function, 220(3), 1355–1368. https://doi.org/10.1007/s00429-014-0729-5

Ising, M., Depping, A.-M., Siebertz, A., Lucae, S., Unschuld, p. G., Kloiber, S., Horstmann, S., Uhr, M., Müller-Myhsok, B., & Holsboer, F. (2008). Polymorphisms in the FKBP5 gene region modulate recovery from psychosocial stress in healthy controls. European Journal of Neuroscience, 28(2), 389–398. https://doi.org/10.1111/j.1460-9568.2008.06332.x

Jacobs, R. H., Barba, A., Gowins, J. R., Klumpp, H., Jenkins, L. M., Mickey, B. J., Ajilore, O., Peciña, M., Sikora, M., Ryan, K. A., Hsu, D. T., Welsh, R. C., Zubieta, J.-K., Phan, K. L., & Langenecker, S. A. (2016). Decoupling of the amygdala to other salience network regions in adolescent-onset recurrent major depressive disorder. Psychological Medicine, 46(5), 1055–1067. https://doi.org/10.1017/S0033291715002615

Jedd, K., Hunt, R. H., Cicchetti, D., Hunt, E., Cowell, R. A., Rogosch, F. A., Toth, S. L., & Thomas, K. M. (2015). Long-term consequences of childhood maltreatment: Altered amygdala functional connectivity. Development and Psychopathology, 27(4 Pt 2), 1577–1589. https://doi.org/10.1017/S0954579415000954

Jenkinson, M., Bannister, P., Brady, M., & Smith, S. (2002). Improved optimization for the robust and accurate linear registration and motion correction of brain images. NeuroImage, 17(2), 825–841.

Jovanovic, T., & Ressler, K. J. (2010). How the neurocircuitry and genetics of fear inhibition may inform our understanding of PTSD. The American Journal of Psychiatry, 167(6), 648–662. https://doi.org/10.1176/appi.ajp.2009.09071074

Kendall-Tackett, K., & Becker-Blease, K. (2004). The importance of retrospective findings in child maltreatment research. Child Abuse & Neglect, 28(7), 723–727. https://doi.org/10.1016/j.chiabu.2004.02.002

Klengel, T., & Binder, E. B. (2015). Epigenetics of Stress-Related Psychiatric Disorders and Gene × Environment Interactions. Neuron, 86(6), 1343–1357. https://doi.org/10.1016/j.neuron.2015.05.036

Klengel, T., Mehta, D., Anacker, C., Rex-Haffner, M., Pruessner, J. C., Pariante, C. M., Pace, T. W. W., Mercer, K. B., Mayberg, H. S., Bradley, B., Nemeroff, C. B., Holsboer, F., Heim, C. M., Ressler, K. J., Rein, T., & Binder, E. B. (2013). Allele-specific FKBP5 DNA demethylation mediates gene-childhood trauma interactions. Nature Neuroscience, 16(1), 33–41. https://doi.org/10.1038/nn.3275

Koch, S. B. J., van Zuiden, M., Nawijn, L., Frijling, J. L., Veltman, D. J., & Olff, M. (2016). Aberrant Resting-State Brain Activity in Posttraumatic Stress Disorder: A Meta-analysis and Systematic Review. Depression and Anxiety, 33(7), 592–605. https://doi.org/10.1002/da.22478

Koopmann, A., Bez, J., Lemenager, T., Hermann, D., Dinter, C., Reinhard, I., Schuster, R., Wiedemann, K., Winterer, G., & Kiefer, F. (2016). The Effect of Nicotine on HPA Axis Activity in Females is Modulated by the FKBP5 Genotype. Annals of Human Genetics, 80(3), 154–161. https://doi.org/10.1111/ahg.12153

Kurth, F., Zilles, K., Fox, p. T., Laird, A. R., & Eickhoff, S. B. (2010). A link between the systems: Functional differentiation and integration within the human insula revealed by meta-analysis. Brain Structure & Function, 214(5–6), 519–534. https://doi.org/10.1007/s00429-010-0255-z

LeDoux, J. (1998). Fear and the brain: Where have we been, and where are we going? Biological Psychiatry, 44(12), 1229–1238.

LeDoux, J. (2007). The amygdala. Current Biology: CB, 17(20), R868–874. https://doi.org/10.1016/j.cub.2007.08.005

Leppänen, J. M. (2006). Emotional information processing in mood disorders: A review of behavioral and neuroimaging findings. Current Opinion in Psychiatry, 19(1), 34–39. https://doi.org/10.1097/01.yco.0000191500.46411.00

Lett, T. A., Waller, L., Tost, H., Veer, I. M., Nazeri, A., Erk, S., Brandl, E. J., Charlet, K., Beck, A., Vollstädt-Klein, S., Jorde, A., Kiefer, F., Heinz, A., Meyer-Lindenberg, A., Chakravarty, M. M., & Walter, H. (2017). Cortical surface-based threshold-free cluster enhancement and cortexwise mediation. Human Brain Mapping, 38(6), 2795–2807. https://doi.org/10.1002/hbm.23563

Lett, T. A., Vogel, B. O., Ripke, S., Wackerhagen, C., Erk, S., Awasthi, S., Trubetskoy, V., Brandl, E. J., Mohnke, S., Veer, I. M., Nöthen, M. M., Rietschel, M., Degenhardt, F., Romanczuk-Seiferth, N., Witt, S. H., Banaschewski, T., Bokde, A. L. W., Büchel, C., Quinlan, E. B., …; IMAGEN Consortium (2020). Cortical Surfaces Mediate the Relationship Between Polygenic Scores for Intelligence and General Intelligence. Cerebral Cortex, 30(4), 2708–2719. https://doi.org/10.1093/cercor/bhz270

Matosin, N., Halldorsdottir, T., & Binder, E. B. (2018). Understanding the Molecular Mechanisms Underpinning Gene by Environment Interactions in Psychiatric Disorders: The FKBP5 Model. Biological Psychiatry, 83(10), 821–830. https://doi.org/10.1016/j.biopsych.2018.01.021

Matsudaira, I., Oba, K., Takeuchi, H., Sekiguchi, A., Tomita, H., Taki, Y., & Kawashima, R. (2019). Rs1360780 of the FKBP5 gene modulates the association between maternal acceptance and regional gray matter volume in the thalamus in children and adolescents. PloS One, 14(8), e0221768. https://doi.org/10.1371/journal.pone.0221768

McCrory, E., Brito, S. A. D., & Viding, E. (2010). Research Review: The neurobiology and genetics of maltreatment and adversity. Journal of Child Psychology and Psychiatry, 51(10), 1079–1095. https://doi.org/10.1111/j.1469-7610.2010.02271.x

McCrory, E., De Brito, S. A., & Viding, E. (2012). The link between child abuse and psychopathology: A review of neurobiological and genetic research. Journal of the Royal Society of Medicine, 105(4), 151–156. https://doi.org/10.1258/jrsm.2011.110222

Medford, N., & Critchley, H. D. (2010). Conjoint activity of anterior insular and anterior cingulate cortex: Awareness and response. Brain Structure & Function, 214(5–6), 535–549. https://doi.org/10.1007/s00429-010-0265-x

Menon, V. (2011). Large-scale brain networks and psychopathology: A unifying triple network model. Trends in Cognitive Sciences, 15(10), 483–506. https://doi.org/10.1016/j.tics.2011.08.003

Menon, V., & Uddin, L. Q. (2010). Saliency, switching, attention and control: A network model of insula function. Brain Structure & Function, 214(5–6), 655–667. https://doi.org/10.1007/s00429-010-0262-0

Mulders, p. C., van Eijndhoven, p. F., Schene, A. H., Beckmann, C. F., & Tendolkar, I. (2015). Resting-state functional connectivity in major depressive disorder: A review. Neuroscience and Biobehavioral Reviews, 56, 330–344. https://doi.org/10.1016/j.neubiorev.2015.07.014

Nicholson, A. A., Sapru, I., Densmore, M., Frewen, p. A., Neufeld, R. W. J., Théberge, J., McKinnon, M. C., & Lanius, R. A. (2016). Unique insula subregion resting-state functional connectivity with amygdala complexes in posttraumatic stress disorder and its dissociative subtype. Psychiatry Research: Neuroimaging, 250, 61–72. https://doi.org/10.1016/j.pscychresns.2016.02.002

Ochsner, K. N., & Gross, J. J. (2005). The cognitive control of emotion. Trends in Cognitive Sciences, 9(5), 242–249. https://doi.org/10.1016/j.tics.2005.03.010

Pagliaccio, D., Luby, J. L., Bogdan, R., Agrawal, A., Gaffrey, M. S., Belden, A. C., Botteron, K. N., Harms, M. P., & Barch, D. M. (2015). Amygdala functional connectivity, HPA axis genetic variation, and life stress in children and relations to anxiety and emotion regulation. Journal of Abnormal Psychology, 124(4), 817–833. https://doi.org/10.1037/abn0000094

Pannekoek, J. N., van der Werff, S. J. A., Meens, p. H. F., van den Bulk, B. G., Jolles, D. D., Veer, I. M., van Lang, N. D. J., Rombouts, S. A. R. B., van der Wee, N. J. A., & Vermeiren, R. R. J. M. (2014). Aberrant resting-state functional connectivity in limbic and salience networks in treatment—Naïve clinically depressed adolescents. Journal of Child Psychology and Psychiatry, and Allied Disciplines, 55(12), 1317–1327. https://doi.org/10.1111/jcpp.12266

Pariante, C. M. (2004). Glucocorticoid receptor function in vitro in patients with major depression. Stress (Amsterdam, Netherlands), 7(4), 209–219. https://doi.org/10.1080/10253890500069650

Phelps, E. A., & LeDoux, J. E. (2005). Contributions of the Amygdala to Emotion Processing: From Animal Models to Human Behavior. Neuron, 48(2), 175–187. https://doi.org/10.1016/j.neuron.2005.09.025

Price, J. L., & Drevets, W. C. (2010). Neurocircuitry of mood disorders. Neuropsychopharmacology: Official Publication of the American College of Neuropsychopharmacology, 35(1), 192–216. https://doi.org/10.1038/npp.2009.104

Pruim, R. H. R., Mennes, M., van Rooij, D., Llera, A., Buitelaar, J. K., & Beckmann, C. F. (2015). ICA-AROMA: A robust ICA-based strategy for removing motion artifacts from fMRI data. NeuroImage, 112, 267–277. https://doi.org/10.1016/j.neuroimage.2015.02.064

Rabinak, C. A., Angstadt, M., Welsh, R. C., Kenndy, A. E., Lyubkin, M., Martis, B., & Phan, K. L. (2011). Altered amygdala resting-state functional connectivity in post-traumatic stress disorder. Frontiers in Psychiatry, 2, 62. https://doi.org/10.3389/fpsyt.2011.00062

Rasetti, R., & Weinberger, D. R. (2011). Intermediate phenotypes in psychiatric disorders. Current Opinion in Genetics & Development, 21(3), 340–348. https://doi.org/10.1016/j.gde.2011.02.003

Reul, J. M., & de Kloet, E. R. (1985). Two receptor systems for corticosterone in rat brain: Microdistribution and differential occupation. Endocrinology, 117(6), 2505–2511. https://doi.org/10.1210/endo-117-6-2505

Roy, A. K., Fudge, J. L., Kelly, C., Perry, J. S. A., Daniele, T., Carlisi, C., Benson, B., Castellanos, F. X., Milham, M. P., Pine, D. S., & Ernst, M. (2013). Intrinsic functional connectivity of amygdala-based networks in adolescent generalized anxiety disorder. Journal of the American Academy of Child and Adolescent Psychiatry, 52(3), 290-299.e2. https://doi.org/10.1016/j.jaac.2012.12.010

Rutter, M., Moffitt, T. E., & Caspi, A. (2006). Gene-environment interplay and psychopathology: Multiple varieties but real effects. Journal of Child Psychology and Psychiatry, and Allied Disciplines, 47(3–4), 226–261. https://doi.org/10.1111/j.1469-7610.2005.01557.x

Scharf, S. H., Liebl, C., Binder, E. B., Schmidt, M. V., & Müller, M. B. (2011). Expression and Regulation of the Fkbp5 Gene in the Adult Mouse Brain. PLOS ONE, 6(2), e16883. https://doi.org/10.1371/journal.pone.0016883

Schumann, G., Loth, E., Banaschewski, T., Barbot, A., Barker, G., Büchel, C., Conrod, p. J., Dalley, J. W., Flor, H., Gallinat, J., Garavan, H., Heinz, A., Itterman, B., Lathrop, M., Mallik, C., Mann, K., Martinot, J.-L., Paus, T., Poline, J.-B., … IMAGEN consortium. (2010). The IMAGEN study: Reinforcement-related behaviour in normal brain function and psychopathology. Molecular Psychiatry, 15(12), 1128–1139. https://doi.org/10.1038/mp.2010.4

Seeley, W. W., Menon, V., Schatzberg, A. F., Keller, J., Glover, G. H., Kenna, H., Reiss, A. L., & Greicius, M. D. (2007). Dissociable intrinsic connectivity networks for salience processing and executive control. The Journal of Neuroscience: The Official Journal of the Society for Neuroscience, 27(9), 2349–2356. https://doi.org/10.1523/JNEUROSCI.5587-06.2007

Shaffer, A., Huston, L., & Egeland, B. (2008). Identification of child maltreatment using prospective and self-report methodologies: A comparison of maltreatment incidence and relation to later psychopathology. Child Abuse & Neglect, 32(7), 682–692. https://doi.org/10.1016/j.chiabu.2007.09.010

Shin, L. M., Wright, C. I., Cannistraro, p. A., Wedig, M. M., McMullin, K., Martis, B., Macklin, M. L., Lasko, N. B., Cavanagh, S. R., Krangel, T. S., Orr, S. P., Pitman, R. K., Whalen, p. J., & Rauch, S. L. (2005). A functional magnetic resonance imaging study of amygdala and medial prefrontal cortex responses to overtly presented fearful faces in posttraumatic stress disorder. Archives of General Psychiatry, 62(3), 273–281. https://doi.org/10.1001/archpsyc.62.3.273

Smith, S. M. (2002). Fast robust automated brain extraction. Human Brain Mapping, 17(3), 143–155. https://doi.org/10.1002/hbm.10062

Smith, S. M., & Nichols, T. E. (2009). Threshold-free cluster enhancement: Addressing problems of smoothing, threshold dependence and localisation in cluster inference. NeuroImage, 44(1), 83–98. https://doi.org/10.1016/j.neuroimage.2008.03.061

Sripada, R. K., King, A. P., Garfinkel, S. N., Wang, X., Sripada, C. S., Welsh, R. C., & Liberzon, I. (2012). Altered resting-state amygdala functional connectivity in men with posttraumatic stress disorder. Journal of Psychiatry & Neuroscience: JPN, 37(4), 241–249. https://doi.org/10.1503/jpn.110069

Teicher, M. H., Samson, J. A., Anderson, C. M., & Ohashi, K. (2016). The effects of childhood maltreatment on brain structure, function and connectivity. Nature Reviews. Neuroscience, 17(10), 652–666. https://doi.org/10.1038/nrn.2016.111

Thomason, M. E., Marusak, H. A., Tocco, M. A., Vila, A. M., McGarragle, O., & Rosenberg, D. R. (2015). Altered amygdala connectivity in urban youth exposed to trauma. Social Cognitive and Affective Neuroscience, 10(11), 1460–1468. https://doi.org/10.1093/scan/nsv030

Tsigos, C., & Chrousos, G. P. (2002). Hypothalamic-pituitary-adrenal axis, neuroendocrine factors and stress. Journal of Psychosomatic Research, 53(4), 865–871.

Uddin, L. Q., Nomi, J. S., Hébert-Seropian, B., Ghaziri, J., & Boucher, O. (2017). Structure and Function of the Human Insula. Journal of Clinical Neurophysiology: Official Publication of the American Electroencephalographic Society, 34(4), 300–306. https://doi.org/10.1097/WNP.0000000000000377

Urry, H. L. (2006). Amygdala and Ventromedial Prefrontal Cortex Are Inversely Coupled during Regulation of Negative Affect and Predict the Diurnal Pattern of Cortisol Secretion among Older Adults. Journal of Neuroscience, 26(16), 4415–4425. https://doi.org/10.1523/JNEUROSCI.3215-05.2006

van der Werff, S. J. A., Pannekoek, J. N., Veer, I. M., van Tol, M.-J., Aleman, A., Veltman, D. J., Zitman, F. G., Rombouts, S. a. R. B., Elzinga, B. M., & van der Wee, N. J. A. (2013). Resting-state functional connectivity in adults with childhood emotional maltreatment. Psychological Medicine, 43(9), 1825–1836. https://doi.org/10.1017/S0033291712002942

VanZomeren-Dohm, A. A., Pitula, C. E., Koss, K. J., Thomas, K., & Gunnar, M. R. (2015). FKBP5 moderation of depressive symptoms in peer victimized, post-institutionalized children. Psychoneuroendocrinology, 51, 426–430. https://doi.org/10.1016/j.psyneuen.2014.10.003

Veer, I. M., Beckmann, C. F., van Tol, M.-J., Ferrarini, L., Milles, J., Veltman, D. J., Aleman, A., van Buchem, M. A., van der Wee, N. J., & Rombouts, S. A. R. B. (2010). Whole brain resting-state analysis reveals decreased functional connectivity in major depression. Frontiers in Systems Neuroscience, 4. https://doi.org/10.3389/fnsys.2010.00041

Veer, I. M., Oei, N. Y. L., Spinhoven, P., van Buchem, M. A., Elzinga, B. M., & Rombouts, S. A. R. B. (2012). Endogenous cortisol is associated with functional connectivity between the amygdala and medial prefrontal cortex. Psychoneuroendocrinology, 37(7), 1039–1047. https://doi.org/10.1016/j.psyneuen.2011.12.001

Wang, Q., Shelton, R. C., & Dwivedi, Y. (2018). Interaction between early-life stress and FKBP5 gene variants in major depressive disorder and post-traumatic stress disorder: A systematic review and meta-analysis. Journal of Affective Disorders, 225, 422–428. https://doi.org/10.1016/j.jad.2017.08.066

White, M. G., Bogdan, R., Fisher, p. M., Muñoz, K. E., Williamson, D. E., & Hariri, A. R. (2012). FKBP5 and emotional neglect interact to predict individual differences in amygdala reactivity. Genes, Brain, and Behavior, 11(7), 869–878. https://doi.org/10.1111/j.1601-183X.2012.00837.x

Wingenfeld, K., Spitzer, C., Mensebach, C., Grabe, H. J., Hill, A., Gast, U., Schlosser, N., Höpp, H., Beblo, T., & Driessen, M. (2010). [The German version of the Childhood Trauma Questionnaire (CTQ): Preliminary psychometric properties]. Psychotherapie, Psychosomatik, Medizinische Psychologie, 60(11), 442–450. https://doi.org/10.1055/s-0030-1247564

Wochnik, G. M., Rüegg, J., Abel, G. A., Schmidt, U., Holsboer, F., & Rein, T. (2005). FK506-binding proteins 51 and 52 differentially regulate dynein interaction and nuclear translocation of the glucocorticoid receptor in mammalian cells. The Journal of Biological Chemistry, 280(6), 4609–4616. https://doi.org/10.1074/jbc.M407498200

Zhang, Y., Brady, M., & Smith, S. (2001). Segmentation of brain MR images through a hidden Markov random field model and the expectation-maximization algorithm. IEEE Transactions on Medical Imaging, 20(1), 45–57. https://doi.org/10.1109/42.906424

