## Supplementary figures and images for "The Interaction of Child Abuse and rs1360780 of the FKBP5 Gene is Associated with Amygdala Resting-State Functional Connectivity in Young Adults"

### Supplemental Figure 1

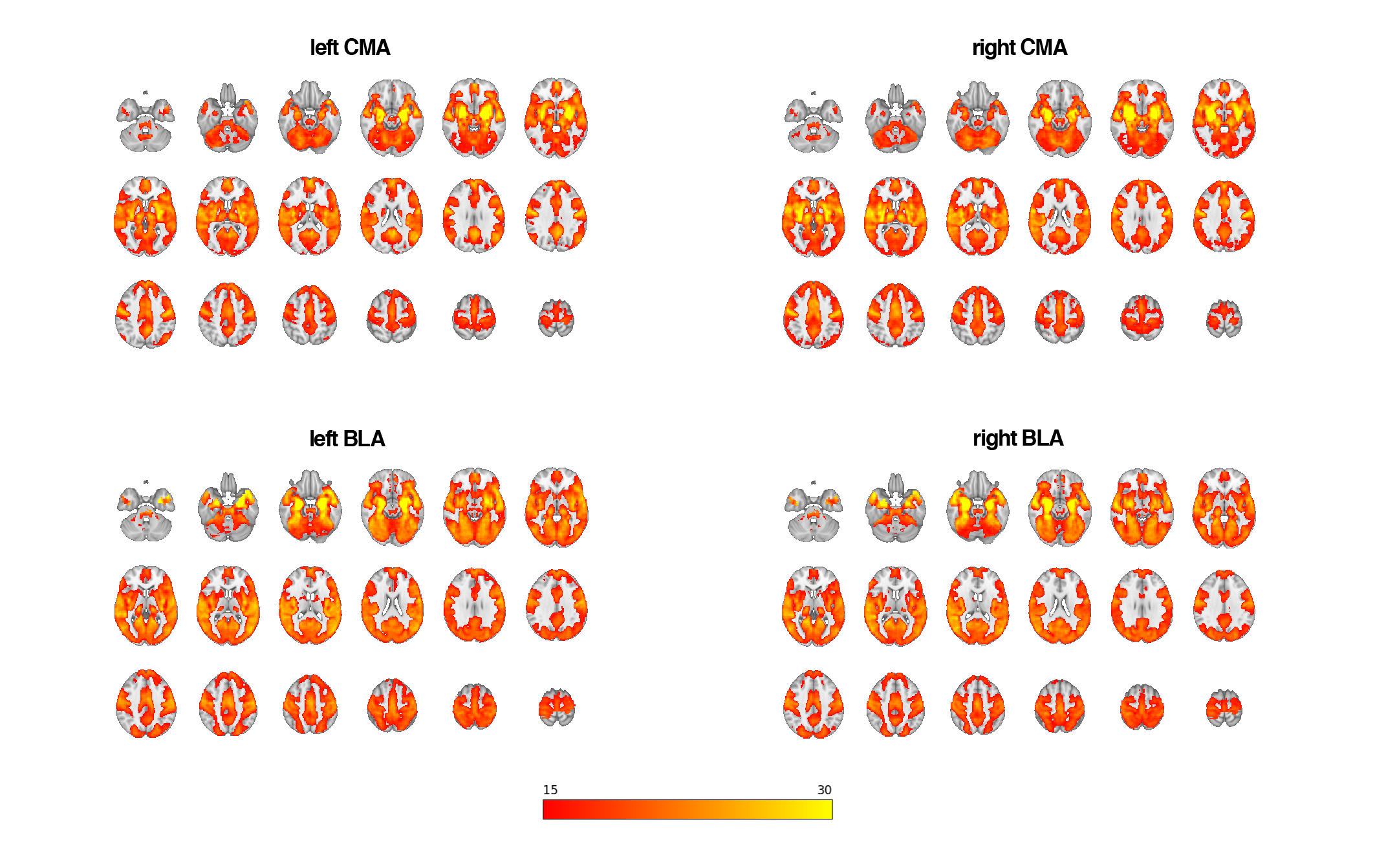
